# tDCS to the left DLPFC improves cognitive control but not action cancellation in patients with ADHD: a behavioral and electrophysiological study

**DOI:** 10.1101/2020.01.13.20017335

**Authors:** Laura Dubreuil-Vall, Federico Gomez-Bernal, Ana C. Villegas, Patricia Cirillo, Craig Surman, Giulio Ruffini, Alik S. Widge, Joan A. Camprodon

**Author notes:** Corresponding author: Laura Dubreuil-Vall, Laboratory for Neuropsychiatry and Neuromodulation, Massachusetts General Hospital, 149 13^th^ St. 10^th^ floor., Boston, MA 02129.

## Abstract

ADHD is a neurodevelopmental disorder associated with significant morbidity and mortality that affects 5% of children, adolescents and adults worldwide. Pharmacological and behavioral therapies exist, but critical symptoms such as dysexecutive deficits remain unaffected. In this randomized, placebo-controlled, double-blind, cross-over study we assess the cognitive and physiological effects of tDCS in adult ADHD patients in order to understand (1) the role of DLPFC laterality in ADHD dysexecutive deficits, (2) the physiological dynamics sustaining the modulation of executive function by tDCS, and (3) the impact of state-dependent dynamics of tDCS effect. The participants consisted of a random sample of 40 adult patients with a primary diagnosis of ADHD. Each patient performed three experimental sessions in which they received 30 minutes of 2mA tDCS stimulation targeting the left DLPFC (anodal F3, cathodal Fp2), the right DLPFC (anodal F4, cathodal Fp1) and Sham. Before and after each session, half of the patients completed the Flanker task (EFT) and the other half performed the Stop Signal Task (SST), while we assessed cognitive (reaction time, accuracy) and neurophysiological measures (EEG Event-Related-Potentials). Results show that anodal tDCS to the left DLPFC modulated cognitive (reaction time) and physiological measures (P300 amplitude) in the Flanker task in a state-dependent manner, but there were no significant effects in the Stop Signal Reaction Time of the SST. We interpret these results as an improvement in interference cognitive control (captured by the EFT) but not in action cancellation (assessed by the SST), supporting the hypothesis of the existence of different impulsivity constructs with overlapping but distinct anatomical substrates and therapeutic strategies. We conclude that anodal tDCS over the left DLPFC using a simple bipolar montage has pro-cognitive effects in dysexecutive patients with ADHD associated with the modulation of physiological signatures of cognitive control (i.e. treatment target), supporting specific hypotheses and strategies for neuromodulation treatment development under an experimental therapeutics framework aiming to link target engagement (cognitive and physiological) with clinical benefit. In addition, we also provide empirical evidence supporting the value of the P200, N200 and P300 as cross-sectional biomarkers of cognitive performance across tasks. Last, we provide mechanistic support for the state-dependent nature of the effects of tDCS, highlighting the importance of controlling the neural states before and during stimulation as a relevant therapeutic strategy.

## 1. Introduction

Attention-deficit hyperactivity disorder (ADHD) is associated with functional impairment and high morbidity and mortality in youth and adulthood^1–3^. Epidemiologic studies suggest that ADHD affects up to 5% of adults worldwide^4–6^. Pharmacological and behavioral therapies exist, but critical symptoms such as dysexecutive deficits persist despite current treatments^7–9^.

Transcranial Direct Current Stimulation (tDCS) is emerging as a promising tool in human neuroscience research and neuropsychiatric therapeutics^10^. Previous studies show that tDCS targeting the dorsolateral prefrontal cortex (DLPFC) modulates domains of executive function^11,12^, specifically those affected in ADHD^13^. None of these studies, however, combined behavioral and physiological measures.

The Eriksen Flanker Task (EFT)^14^ and the Stop Signal Task (SST)^15^ are well-established experimental tasks used to assess impaired executive functions in ADHD^16,17^. Although they both capture inhibitory control processes, the EFT primarily assesses interference cognitive control (the ability to resist or resolve distracting interference that is irrelevant to the task), while the SST measures action cancellation (the ability to suppress dominant, automatic, already initiated responses)^18–20^. Human electrophysiological studies assessing event related potentials (ERPs) have established relevant signatures of executive function during these tasks. Specifically, P200, N200, P300 and ERN/Pe (Error Related Negativity/Positivity) characterize the attentional and inhibitory functions that break down during conflict due to dysexecutive deficits and impulsivity^21–24^.

In this study, we tested 40 ADHD patients during three experimental visits and compared the effect of anodal tDCS targeting the left DLPFC vs. right DLPFC vs. Sham. Immediately before and after tDCS, half of the patients performed the EFT and the other half the SST, while we measured behavioral (reaction time and accuracy) and neurophysiological (ERPs) responses. The primary goal of the study was to empirically validate physiological biomarkers of dysexecutive deficits in adult ADHD, including response biomarkers (i.e. therapeutic targets), cross-sectional diagnostic biomarkers) and physiological and behavioral baseline prognostic signatures characterizing state-dependent effects^25^. Our specific aims were to assess (1) the role of DLPFC laterality in ADHD deficits in interference cognitive control (EFT) and action cancellation (SST), (2) the physiological dynamics sustaining the modulation of executive function by tDCS and (3) the impact of state-dependent dynamics of tDCS effects.

## 2. Methods and materials

### 2.1 Trial design

A randomized, placebo-controlled, double-blind, cross-over study was performed at the Massachusetts General Hospital (Boston, USA). Recruitment started on July 2015 and ended on March 2018. The study was approved by the Partners Healthcare Institutional Review Board and registered on ClinicalTrials.gov (NCT04175028). The full protocol is available at Supplement 1.

### 2.2 Participants

Forty-four adult patients with a primary diagnosis of ADHD were recruited from the Division of Neuropsychiatry, the Behavioral Neurology Unit and the Adult ADHD Research Program at Massachusetts General Hospital and randomized (see Table S1 in Supplement 2 for demographic characteristics and medications). Patients between 18 and 67 years of age with a diagnosis of ADHD in adulthood were included, meeting the DSM-5^26^ criteria determined by semi-structured clinical interview (MINI International Neuropsychiatric Interview) and clinical questionnaires (Adult Self-Reported Scale, ASRS). Patients were either off stimulant medications or, if undergoing treatment with stimulants, were asked to discontinue two days prior to the experiment, under physician-guided protocol, and allowed to resume afterwards. Exclusion criteria consisted of current or past history of clinically unstable psychiatric conditions, pregnant or nursing females, and inability to participate in testing procedures or contraindications to tDCS. All participants gave informed and written consent for participation.

### 2.3 Intervention

For each session, 2mA of anodal stimulation was applied for 30 minutes with Ag/AgCl electrodes (contact area 3.14 cm^2^) using the hybrid tDCS-EEG Starstim® system (Neuroelectrics, USA, also used for EEG recording). The duration of the ramp-up and -down at the beginning and the end of the stimulation was set to 15 seconds. During the stimulation period the subject was instructed to sit and relax with eyes open.

The anodal electrode was placed on the scalp at the F4 (for Right DLPFC stimulation) or F3 (for Left DLPFC stimulation) positions, according to the international 10-20 EEG system (Figure S0 in Supplement 2). The cathode was placed in the contralateral supraorbital region (Fp1 or Fp2). For the sham condition, the electrodes were placed at the same positions but the current was applied only for the 15-second ramp up phase at the beginning and the end of a 30-minute sham-stimulation period, to simulate the potential experience of local tingling sensation that real stimulation produces but without sustained effect on cortical activity.

The order of stimulation was randomized by a computer-generated list. The experimenter and the subject were blinded by using the double-blind mode in Starstim’s software NIC, which blinds the user on the type of stimulation used (active tDCS targeting left/right DLPFC or sham) after a 4-digit password is introduced by the administrator.

### 2.4 Outcomes

Immediately before and after tDCS, half of the patients (n=20) completed the EFT (Figure 2a), in which subjects must respond to the direction of a central arrow that is surrounded (“flanked”) by distracting arrows that can either have the same (congruent trials) or opposing orientation (incongruent trials) as the central one. Participants were instructed to press the left or right arrow buttons following the direction of the central arrow, ignoring the flanker arrows. The accuracy of correct/incorrect responses and the reaction time (RT) for each stimulus were measured.

**Figure 1.**
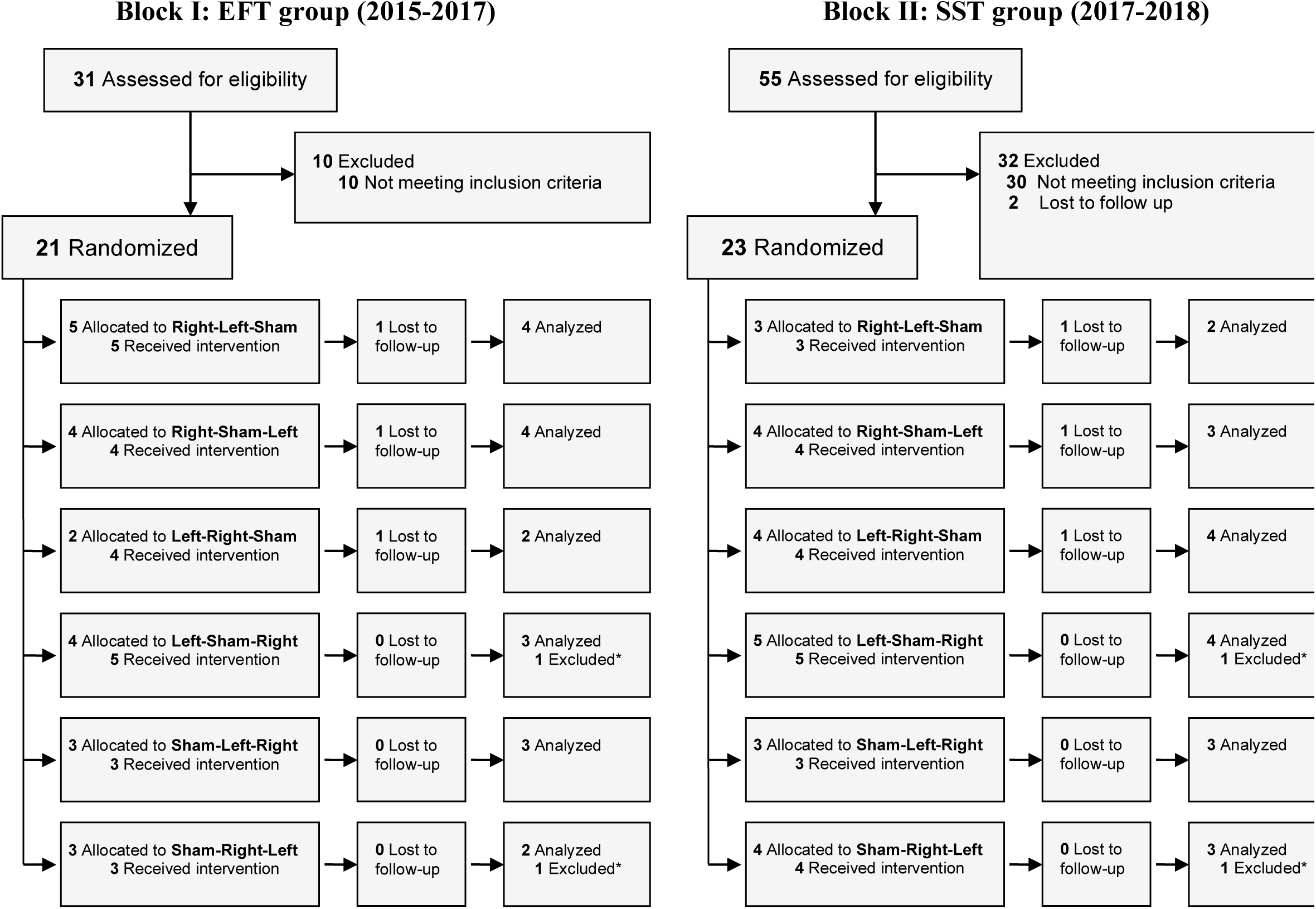
CONSORT Diagram.

**Figure 2.**
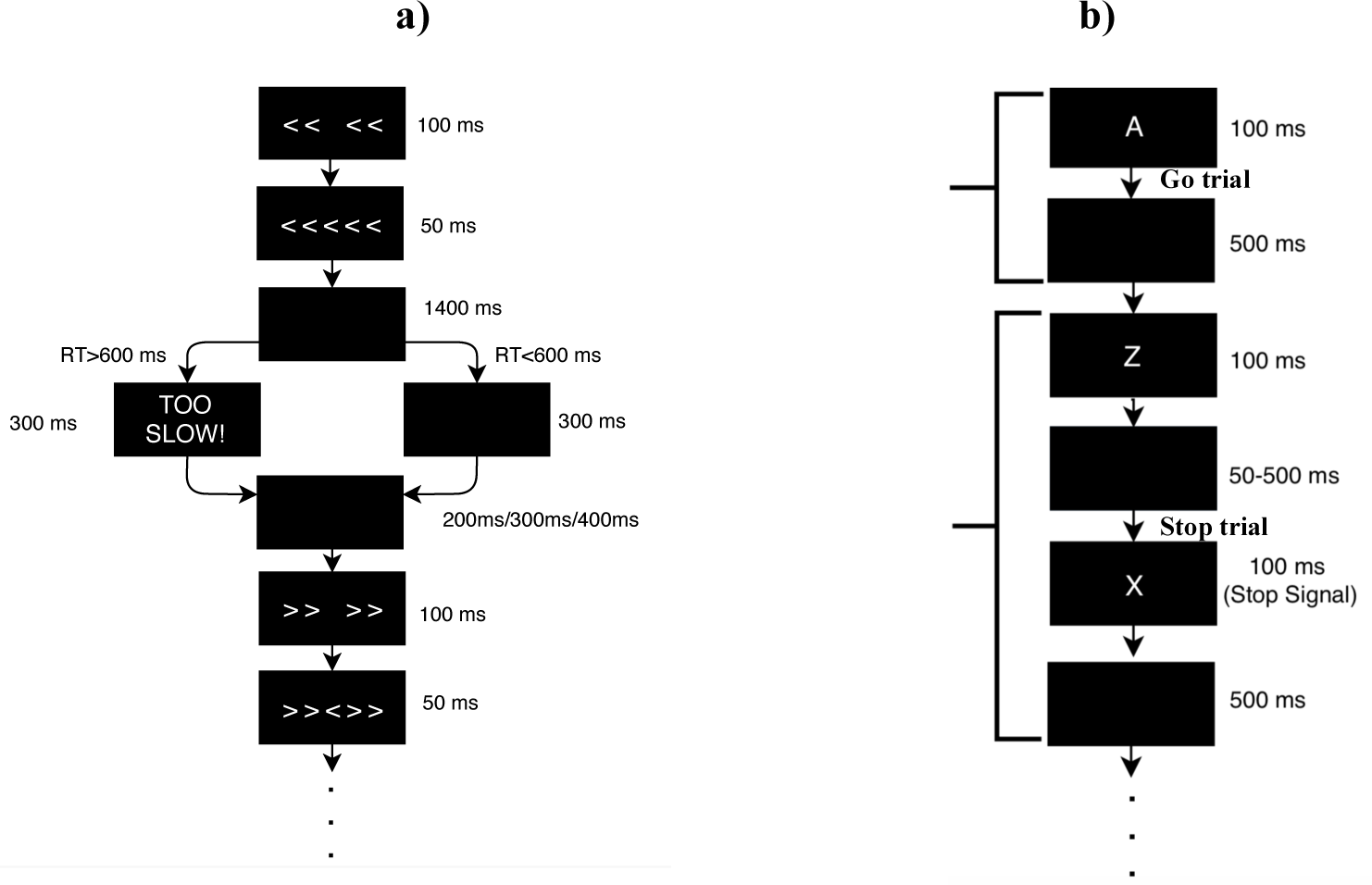
Flanker and Stop Signal task scheme. **a)** The Flanker task consisted of 140 trials in two blocks of 70. Each subject had a different, fully random sequence of congruent and incongruent trials, with 2 congruent trials for each incongruent trial, in order to build a tendency towards congruent responses and thus increase the difficulty of conflict detection in incongruent trials. The task had a total duration of 10 min, with 1 minute of training before the task started. The flanker arrows were first presented alone for duration of 136ms, 114ms, 92ms, 70ms or 48ms, and were then joined by the target arrow for 62ms, 52ms, 42ms, 32ms or 22ms, respectively (values were adjusted to the pyschometric “sweet spot” in which each patient achieved a performance in the range of 80-85%). Stimulus presentation was followed by a black screen for 500 ms. The time-window for participants’ response was 600 ms after target onset. If the participant did not respond within the response window, a screen reading ‘TOO SLOW!’ was presented for 300 ms. Participants were told that if they saw this screen, they should speed up. If a response was made before the deadline, the ‘TOO SLOW!’ screen was omitted, and the black screen remained on screen for the 300 ms interval. Finally, each trial ended with presentation of the fixation cross for an additional randomly chosen duration (200, 300 or 400 ms) in order to avoid any habituation or expectation by the subject. Thus, trial durations varied between 1070–1400 ms. **b)** The SST consisted of 160 Go-trials (80%) and 40 Stop-trials (20%). The “A” or “Z” stimuli were first presented for 100ms and they were followed by a black screen for 500ms. For the Stop-trials, the Stop Signal initially appeared 400ms after the “A” or “Z”, and was adjusted dynamically according to the subject’s performance, increasing or decreasing by 50ms respectively after a successful or unsuccessful answer, within a range of 50 to 500ms in order to yield approximately 50% successful inhibition of the Stop-trials.

The other half of the patients (n=20) performed the SST (Figure 2b), in which participants must press the right or left mouse button as quickly as possible when letters “Z” or “A” appear, respectively (Go-trial). However, whenever “A” or “Z” is followed by an “X” (the Stop Signal), which appears with varying adaptive delays from the Go stimulus, participants must withhold their response (Stop-trial). We measured the accuracy of correct/incorrect responses, the RT for Go-trials and the time it takes for the participant to withhold their response in the Stop-trials (Stop-Signal Reaction Time, SSRT).

During the tasks, EEG was recorded from 7 positions (Fp1, Fp2, F3, Fz, F4, P3 and P4) with Ag/AgCl electrodes at a sampling frequency of 500 samples/second. EEG data was referenced to the right mastoid. Offline processing was then performed using EEGlab (version 13.5.4b)^27^. Independent component analysis (ICA) was utilized to identify and remove activity associated with blinks, eye movements, and other artifacts. Data was filtered from 1 to 20 Hz to remove non-neural physiological activity (skin/sweat potentials) and noise from electrical outlets. Trials were epoched within a time frame of 200 ms before and 800 ms after the stimulus onset. The mean of the pre-stimulus baseline [−200,0] ms was then subtracted from the entire ERP waveform for each epoch to eliminate any voltage offset. After rejecting trials that had at least a sample above +/−150 uV, the remaining trials were averaged for each time point and stimulation condition.

### 2.5 Statistical analysis

Based on similar studies^12,28^, we estimated a sample size of 20 subjects for each block of tasks to provide 85% power to detect an effect size of d=0.6 (α=5%), 25 subjects would provide XX% power and 30 subjects YY % power for the same estimated effect size and α=5%. We estimated a 20% attrition rate based on previous studies.

The RT and the ERP amplitudes were introduced into Generalized Linear Models with Mixed Effects (GLMM) modeled using the *glmer* and *lmer* functions of the *lme4* package^29^ in R software (v1.0.136)^30^ (see Tables S2-S7 in Supplement 2 for expanded details). A Gamma distribution was used for the RT, a binomial distribution was used for the accuracy of correct responses and a Gaussian distribution was used to model the ERP amplitudes (see Supplement 2 for more details). Correction for multiple comparisons was applied using the ‘mvt’ method^31^ from the *lsmeans* package in R, which produces the same adjustments as the Tukey method but using Monte-Carlo simulations^31^. Coefficients were considered significant when p<0.05 (two-tailed).

For the Stop-trials of the SST, the mean SSRT was calculated by subtracting the mean Stop Signal Delay (SSD: the time it takes for the Stop Signal to appear after the “A” or “Z” stimuli) from the mean RT of Go-trials^32^. The mean SSRT was then analyzed using a standard ANOVA model with the Stimulation type (Left/Right/Sham) and the TimePoint (PRE/POST) as factors.

ERP analysis focused on P200, N200 and P300 amplitudes for incongruent EFT trials and SST Stop-trials, and P200 in congruent EFT trials and SST Go-trials (as congruent and Go-trials do not show N200 and P300). ERN and Pe were also analyzed in both tasks for trials with incorrect responses. After visual inspection of the grand average waveforms, the mean ERP amplitude of each single trial was calculated in a specific time-window centered in the peak latency of the grand average waveforms. Mean amplitudes were then introduced into a linear model with mixed effects (LMM) and a normal distribution. ERP analysis focused on the average of F3, Fz and F4 positions.

For state-dependencies, the RT and the amplitude of P200, N200 and P300 were computed separately for real tDCS and Sham. Beta coefficients were modeled with a linear model with mixed effects (LMM), with the difference from PRE to POST stimulation as the dependent variable and the mean of the same (or another) variable at baseline as an independent factor. P values were corrected for multiple comparisons using the False Discovery Rate (FDR) method.

## 3. Results

As shown in Figure 1, 37 patients were included in the analysis: 18 in the EFT group and 19 in the SST group. Attrition rate was lower than anticipated. No important harms or unintended side effects were reported.

### 3.1 Flanker task

#### 3.1.1 Cognitive results

For incongruent trials, left-sided stimulation led to a significantly faster RT compared to Sham (β=−16.1ms, CI=[−22.8, −9.3], p<0.0001), while right-sided stimulation did not have any significant effect (β=−5.3ms, CI=[−13.7, 3.1], p=0.390) (Figure 3a). The effect of left-sided stimulation was also greater than that of right-sided stimulation (β=10.7ms, CI=[1.26, 20.2], p=0.0183). For congruent trials there were no significant changes for any of the stimulation conditions (Figure S1a in Supplement 2). None of the stimulation conditions lead to significant changes in accuracy compared to Sham, both for incongruent and congruent trials (Figure 3b and S1b).

**Figure 3.**
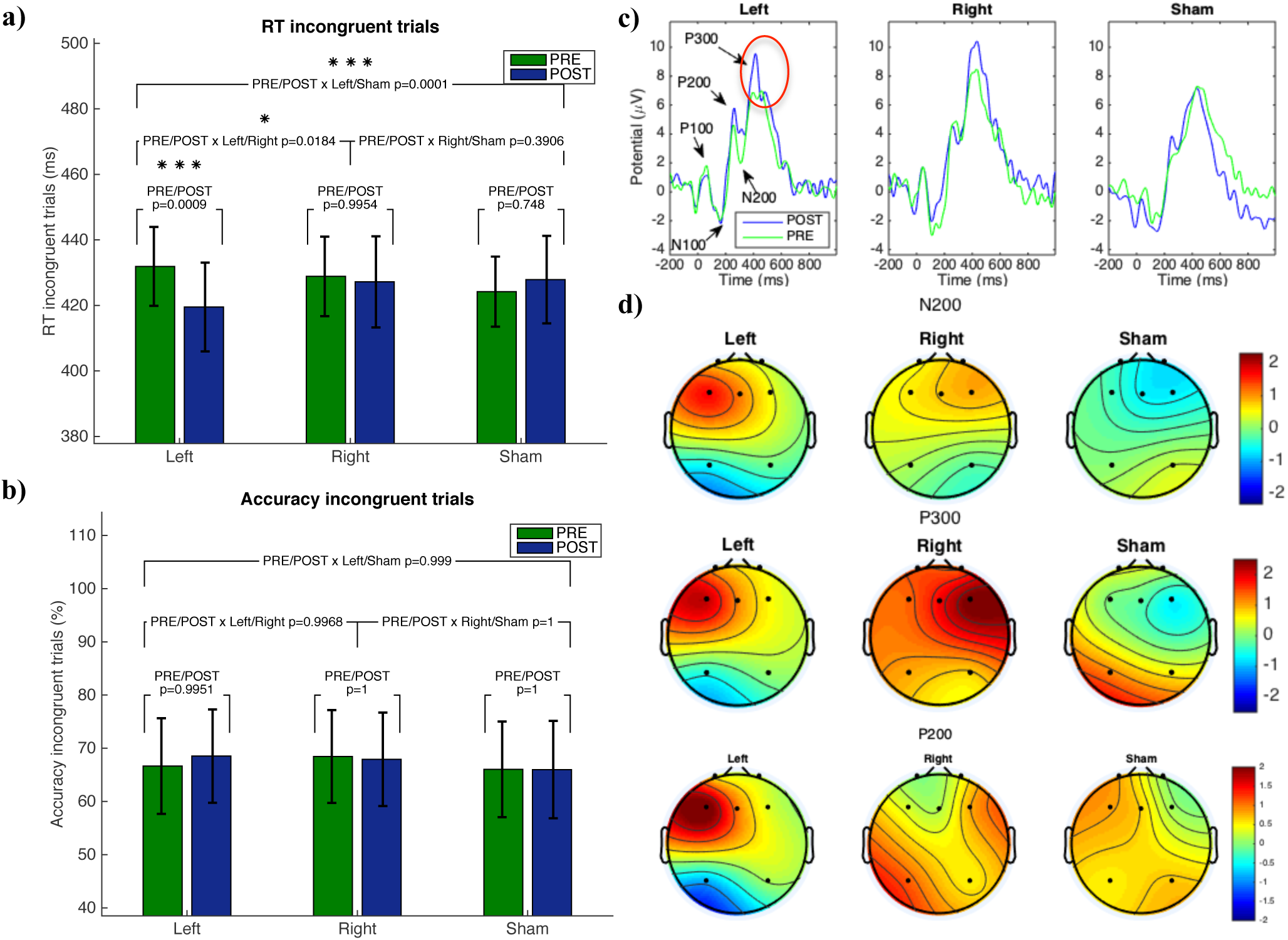
Flanker results. **a)** Mean reaction time and **b)** accuracy for incongruent trials and p values with ‘mvt’ correction. Error bars indicate confidence intervals. Significance indicated as (*) = p<0.05, (**) = p<0.01, (***) = p<0.001. **c)** Grand average ERPs time-locked to incongruent stimuli. Waveforms correspond to the average of F3, Fz and F4 positions. The red circle indicates the significant amplitude changes compared to Sham. See Figure S2 in Supplement 2 for ERPs at individual electrodes. **d)** Scalp topographies of POST-PRE difference of P200, N200 and P300 amplitude (uV). Averaging time window for P300=[260, 360] ms. Averaging time window for N200=[180, 230]ms.

#### 3.1.2 Event-Related Potentials

Both left-sided stimulation (β=2.15 µV, CI=[0.31, 3.99], p=0.022) and right-sided stimulation (β=2.37 µV, CI=[0.53, 4.20], p=0.011) led to a significant P300 amplitude increase compared to Sham for incongruent trials (Figure 3c).

There are no significant changes in N200 after left-sided or right-sided stimulation compared to Sham, but the reduction in N200 amplitude after left-sided stimulation is significantly different compared to right-sided stimulation (β=−2.43 µV, CI=[−4.64, −0.22], p=0.027). Note that most P200, N200 and P300 amplitude changes occurred primarily around the area of the anodal stimulation electrode (F3 or F4), matching the laterality of the stimulation, especially for left-sided stimulation (Figure 3d).

There were no significant changes in P200 amplitude for incongruent (Figure 3b) or congruent (Figure S3a in Supplement 2) trials. ERN and Pe did not show significant differences either (Figure S3b in Supplement 2).

#### 3.1.3 ERP cross-correlation with reaction time

The amplitudes of P200, N200 and P300 were significantly correlated with RT for incongruent trials in a cross-sectional trial-by-trial basis: the greater P200 (β=−0.26ms/µV, CI=[−0.43, −0.09], p=0.002) and P300 amplitudes (β=−0.25ms/µV, CI=[−0.50, −0.04], p=0.046) the faster the RT, and the smaller the N200 amplitude the faster the RT (β=−0.54ms/µV, CI=[−0.79, −0.29], p<0.0001).

#### 3.1.4 State-dependencies

Table S8 in Supplement 2 shows the effect of variables at baseline (before stimulation) on the change on the same (and other) variables after stimulation (i.e. state-dependent relationships). Figure 5 shows the scatter plots of the significant predictors. The change in P300 and P200 after stimulation is conditioned by the amplitude of P300 and P200 at baseline, while the change in N200 is conditioned by the amplitude of N200 and P300 at baseline.

**Figure 5.**
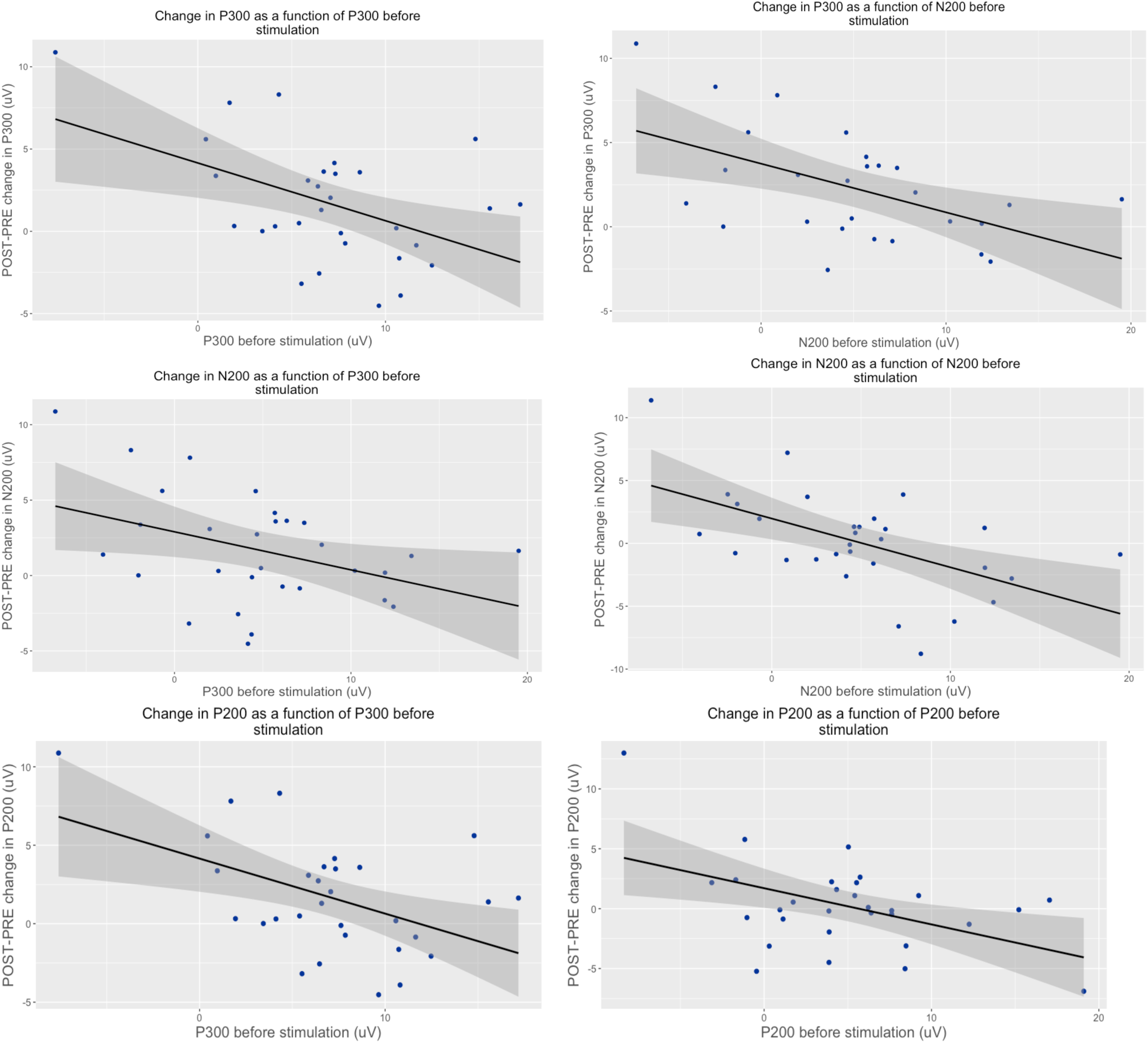
Scatter plots, regression lines and confidence intervals for significant state dependencies in the EFT. **Top row:** change in P300 as a function of P300 (left) and P200 (right) at baseline in the EFT. **Second row:** change in N200 amplitude as a function of P300 (left) and N200 (right) amplitude at baseline in the EFT. **Third row:** change in P200 amplitude as a function of P300 (left) and P200 (right) amplitude at baseline in the EFT.

### 3.2 Stop Signal Task

#### 3.2.1 Cognitive results

The RT for Go-trials significantly increased after Left stimulation compared to sham (β=8.32 µV, CI=[2.18, 14.47], p=0.0044) (Figure 4a), but there were no significant changes in the SSRT for Stop-trials (Figure 4b). There were also no significant changes in accuracy for any of the stimulation conditions and for none of the trial types (Figure S4 in Supplement 2).

**Figure 4.**
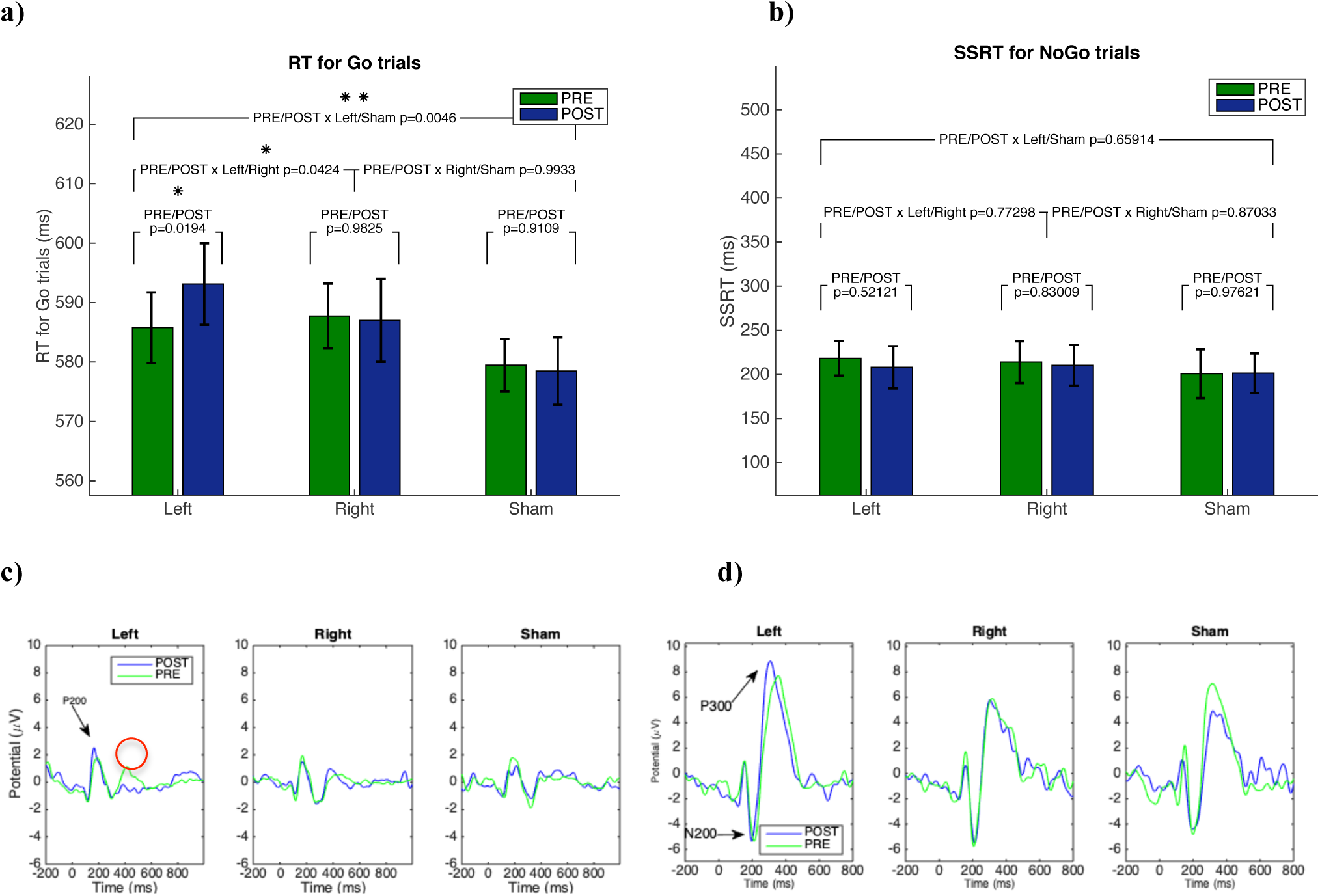
SST results. **a)** Mean reaction time for Go-trials. **b)** SSRT for Stop-trials. Error bars indicate confidence intervals. Significance indicated as (*) = p<0.05, (**) = p<0.01. **c)** Grand average ERPs time-locked to Go-trials. The red circle indicates the significant amplitude changes compared to Sham. **d)** Grand average ERPs time-locked to Stop-trials. Waveforms correspond to the average of F3, Fz and F4 positions.

#### 3.2.2 Event-Related Potentials

For Go-trials, P200 amplitude significantly increased after Left stimulation compared to Sham (β=0.51uV, CI=[0.09, 0.92], p=0.0160) (Figure 4c). For Stop-trials, there were no significant changes in P200, N200 and P300 (Figure 4d). There were no significant changes in ERN or Pe (Figure S5 in Supplement 2).

#### 3.2.3 ERP cross-correlation with reaction time

The amplitude of P200 was also significantly correlated with RT for Go-trials, i.e., the greater P200 amplitude, the slower the RT (β=1.08ms/µV, CI=[0.69, 1.47], p<0.001).

#### 3.2.4 State-dependencies

No significant state-dependencies were found for the SST (Table S8 in Supplement 2).

## 4. Discussion

To the best of our knowledge, this is the first study with ADHD patients to assess the lateralized effects of anodal tDCS to the left and right DLPFC combining cognitive with physiological measures of executive function, using different inhibitory control tasks, quantifying state-dependent dynamics and assessing cross-sectional physiological biomarkers.

### 4.1 Flanker Task

Our results confirmed that anodal tDCS targeting the DLPFC improves RT in the EFT in ADHD patients, similarly to what we previously described in healthy subjects^12^. Specifically, we describe that anodal tDCS targeting the left DLPFC results in a significant reduction in RT in incongruent trials, compared to a non-significant change after sham or right DLPFC anodal modulation. Compared to our previous study with HC, the baseline RT (before stimulation) is slower in ADHD patients and the size effect of the improvement in RT after left-sided stimulation is larger for ADHD (β=−16.1ms) than for HC (β=−8.37ms).

The faster RT of incongruent trials in ADHD patients is correlated with a significant increase in P300 amplitude, in this case for both left- and right-sided anodal tDCS. Larger P300 amplitudes are associated with effective conflict post-processing and cognitive control, with the subsequent behavioral inhibition of incorrect prepotent responses^22,33,34^. We thus interpret the increase in P300 amplitude as a modulation of conflict resolution and interference control processes, leading to more efficient inhibition of distractors and competing responses (i.e., faster RT). Given that P300 increases significantly after left-sided and right-sided stimulation, but behavior (RT) only changes after left-sided stimulation, we hypothesize that the physiological effect of right-sided stimulation is not sufficient to trigger a significant behavioral change, suggesting a greater role for the left DLPFC in the modulation of executive function.

Although in previous research with healthy controls we found a significant decrease in N200 amplitude after left stimulation^12^, indicative of an improvement in selective attention in a context of conflict resolution, in the current study the decrease in N200 amplitude was not significant compared to Sham. This highlights the need to better understand the underlying physiological differences between ADHD patients and healthy subjects leading to different tDCS effects.

### 4.2 Stop Signal Task

In the SST, *proactive inhibition* is defined as the advanced preparation to halt action in the anticipation of an imminent Stop Signal in Go-trials, requiring greater selective attention in the visual search for the Stop Signal to appear. *Reactive inhibition* is defined as the performance of outright stopping in response to the appearance of a Stop Signal in Stop-trials^35^. In the current study we found that tDCS to the left DLPFC leads to a significant increase on the time patients withhold their response in Go-trials waiting for the Stop Signal to appear, which is correlated with a significant increase in P200 amplitude. There is a wide range and diversity of factors that have been found to affect the characteristics of the P200, but its amplitude is generally associated with selective attention to visual stimuli^36^. We thus interpret the increase in P200 amplitude as a modulation in selective attention when searching for the Stop Signal to appear, with the subsequent improvement in proactive inhibition. However, the lack of significant changes in the SSRT suggests no effects on reactive inhibition. Although there have been positive results in tDCS studies using the SST in a healthy population targeting other areas like the right inferior frontal gyrus^37–42^, previous tDCS studies with ADHD patients using the SST and targeting the DLPFC have also found a lack of significant effects on the SSRT and accuracy^28,43^. These results support the formulation of *inhibitory control and impulsivity as complex multimodal processes with subdomain specificity (e*.*g. impulsivity of thought, action, affect, etc*.*) captured by different experimental tasks and with different anatomical representation*^*44*^. These findings confirm the hypothesis that that there is a dissociation between action cancellation or the ability of suppressing prepotent responses that have already been initiated, captured by the SST, and interference cognitive control or the ability of resisting distractors and resolving interference between competing responses, captured by the EFT^20,45–50^. From a translational perspective, this also implies that the DLPFC may be a good substrate to improve interference cognitive control and proactive inhibition, but if the goal is to improve action cancellation (e.g. tics, compulsions) one should consider alternative windows into the circuitry^19,44^.

### 4.3 Cross-sectional biomarkers

Our findings indicate that the amplitude of P200, N200 and P300 on a trial-by-trial basis is correlated cross-sectionally with RT in the EFT (as previously described in healthy subjects^12^) and the SST, thus supporting the interpretation of the observed tDCS effects (i.e. small N200 and large P300 are associated with more adaptive responses thus changes in this direction should be therapeutic) and highlighting their value as a potential diagnostic, monitoring or surrogate biomarker^25^ for cognitive performance.

### 4.4 State-dependence

Smaller N200 and greater P300 amplitudes are associated with better cognitive performance^21,33^, therefore decreases in N200 and increases in P300 can be interpreted as improvements. Our findings indicate that the more maladaptive the baseline ERP amplitude, the greater the improvement after tDCS in the EFT. The state-dependent characteristics of tDCS have important implications for treatment development: beyond stimulation parameters, clinical trials may benefit from controlling patients’ baseline state (before or during stimulation) to minimize response variability and maximize the therapeutic effects of tDCS.

### 4.3 Limitations

While the field has established differences between the constructs captured by the EFT and the SST, the nomenclature used to define the overlap and differences at the cognitive and behavioral level remain equivocal and often contradictory. This seems more than a simple problem with semantics and reflects deficits in the core formulation of the subtleties across executive constructs. We have opted for descriptive terms previously used in the literature (e.g. interference cognitive control and action cancellation), but acknowledge that other terminologies may be considered.

### 4.4 Conclusions

This study indicates that anodal tDCS over the left DLPFC using a simple bipolar montage has pro-cognitive effects in dysexecutive patients with ADHD associated with the modulation of physiological signatures of cognitive control (i.e. treatment target), supporting specific hypotheses and strategies for neuromodulation treatment development under an experimental therapeutics framework aiming to link target engagement (cognitive and physiological) with clinical benefit. In addition, we provide mechanistic support for the state-dependent nature of the effects of tDCS, highlighting the importance of controlling (or at least measuring) the neural states before (and possibly during) stimulation as a relevant therapeutic strategy beyond choices regarding the neuromodulation parameter space. We also provide empirical evidence supporting the value of the P200, N200 and P300 as cross-sectional biomarkers of cognitive performance across tasks, and across populations if taken together with our previous similar report in healthy subjects^12^. Last, we interpret these results as an improvement in interference cognitive control (captured by the EFT) but not in action cancellation (assessed by the SST), supporting the hypothesis of the existence of different impulsivity constructs with overlapping but distinct anatomical substrates and therapeutic strategies.

## Data Availability

Data will be made available upon reasonable request to the senior author (JAC).

## 5. Acknowledgements

This research was partly supported by NIH grants (RO1 MH112737, R21 DA042271, R21 AG056958 and R21 MH115280) to JAC.

## Contributions

JAC designed the study; AV, FGB and PC acquired the data; LDV, GR, ASW, JAC analyzed the data; LDV, CS, ASW and JAC wrote the manuscript.

